# SARS-CoV-2 detection in nasopharyngeal throat swabs by metagenomics

**DOI:** 10.1101/2020.05.24.20110205

**Authors:** Le Van Tan, Nguyen Thi Thu Hong, Nghiem My Ngoc, Tran Tan Thanh, Vo Thanh Lam, Lam Anh Nguyet, Le Nguyen Truc Nhu, Nguyen Thi Han Ny, Ngo Ngoc Quang Minh, Dinh Nguyen Huy Man, Vu Thi Ty Hang, Phan Nguyen Quoc Khanh, Tran Chanh Xuan, Nguyen Thanh Phong, Tran Nguyen Hoang Tu, Tran Tinh Hien, Le Manh Hung, Nguyen Thanh Truong, Lam Minh Yen, Nguyen Thanh Dung, Guy Thwaites, Nguyen Van Vinh Chau, for OUCRU COVID-19 research group

## Abstract

Metagenomics could detect SARS-CoV-2 in all eight nasopharyngeal/throat swabs with high/low viral loads, and rhinovirus in a co-infected patient. The sequenced viruses belonged to lineage B1. Because metagenomics could detect novel pathogen and co-infection, and generate sequence data for epidemiological investigation, it is an attractive approach for infectious-disease diagnosis.

---

Metagenomics is a sensitive sequence-independence method for infectious disease diagnosis and the discovery of novel pathogens [1]. The novel coronavirus namely severe acute respiratory syndrome coronavirus 2 (SARS-CoV-2) is the cause of the ongoing coronavirus disease 2019 (COVID-19) pandemic [2]. However, there have only been three studies reporting the utility potential of metagenomics to detect SARS-CoV-2 directly from clinical specimens, with a combined sample size of nine patients [3-5]. But none of these has been conducted in resource-limited settings. In this area of the world, emerging infection however is likely to emerge. Here we describe the application of metagenomics to detect SARS-CoV-2 in RT-PCR positive nasopharyngeal throat swabs. In addition, using the obtained sequence, we genetically characterize the viruses.

## THE STUDY

Since the beginning of March, 2020 an observational study have been conducted at the Hospital for Tropical Diseases (HTD) in Ho Chi Minh City, Vietnam and another one at one of its two designated centres for receiving and treating COVI-19 patients from southern Vietnam with a population of over 40 million (Figure 1). We enrolled patients with a confirmed SARS-CoV-2 diagnosis admitted to the study settings within 48 hours. We collected nasopharyngeal throat swabs (NTS), clinical and laboratory data, and travel and contact history from each study participant. The collected NTS was stored at 4°C at the study sites within four hours and was then transferred to the clinical laboratory of HTD for analysis. SARS-CoV-2 detection was carried out using a WHO recommended real time RT-PCR assays [6]. Assessment of co-infection with common respiratory viruses was carried out using multiplex RT-PCR targeted at 15 different respiratory viruses [7]. The clinical studies received approvals from the Institutional Review Board of the HTD and the Oxford Tropical Research Ethics Committee of the University of Oxford. Study participants gave their written informed consent.

**Figure 1:**
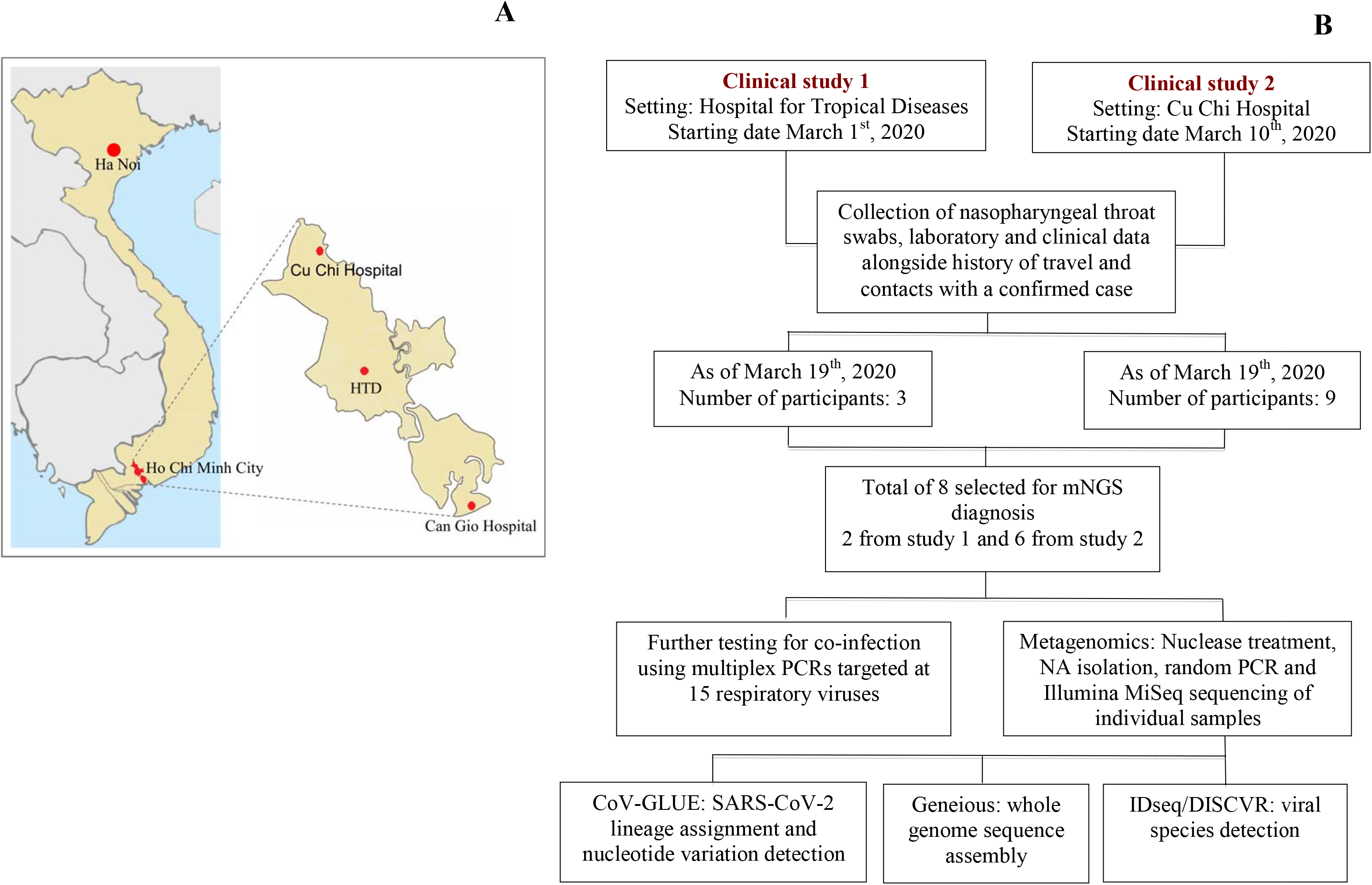
Flowchart showing the study settings and the studies. (**A**) Map showing the location of the for Tropical Diseases (HTD) main campus and its ed COVID-19 centres in Cu Chi where the clinical is conducted, and Can Gio, (**B**) Flowchart ng the patient enrolment, mNGS analysis and post ysis of the obtained sequences **Note to Figure 1**: Maps were obtained from https://mapchart.net/.

The selected samples were individually analyzed with the inclusion of a molecular grade water sample serving as a non-template control (NTC). Metagenomics was carried out as previously described [8]. DNA libraries of individual samples and NTC were then multiplexed using double unique indexes (i.e. each sample was differentiated by double barcodes) and sequenced on an Illumina MiSeq platform using a 300-cycle MiSeq reagent kit V3 (Illumina). Detection of SARS-CoV-2 and co-infection viruses in the obtained sequence data was carried out using a combination of publically availably metagenomics pipelines namely IDseq (idseq.net) and DISCVR [9]. Reference based mapping approach was applied to assemble SARS-CoV-2 genomes from the obtained sequences using Geneious 11.0.3 (Biomatters, Auckland, New Zealand). SARS-CoV-2 lineage determination and detections of nonsynonymous mutations were carried out using CoV-GLUE (http://covglue.cvr.gla.ac.uk), a publically available tool for SARS-CoV-2 sequence analysis (Figure 1).

As of March 19^th^, 2020, a total of 11 PCR confirmed SARS-CoV-2 patients were enrolled in the clinical studies (Figure 1). As a pilot, we selected eight with a wide range of viral loads, as reflected by real time Cycle threshold (Ct) values, for metagenomics analysis (Figure 2A). Information about demographics and clinical status of the eight included patients are presented in Table 1. All were adults and two were asymptomatic carriers identified through contact tracing approach implemented in Vietnam [10]. Three were cases of locally acquired infection and five were imported cases, and one was co-infected with rhihnovirus.

**Figure 2:**
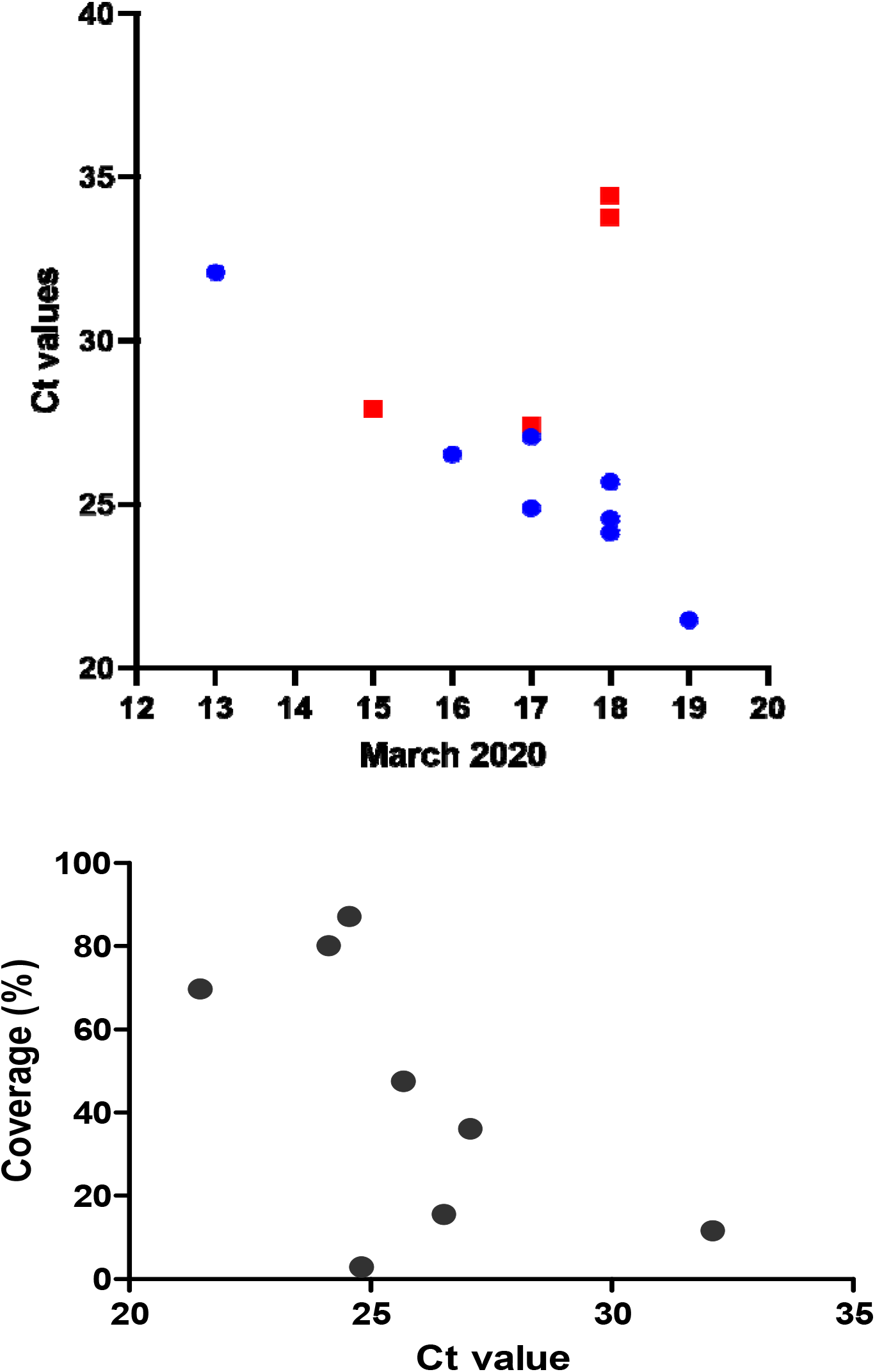
Distribution of Ct values of nasopharyngeal throat swabs of the study participant (A), and s association between Ct values and genome coverage of SARS-CoV-2 generated by mNGS (B) **Note to Figure 2A**: Blue dot represent for samples selected for mNGS while red squares represent for samples not selected for mNGS. Numbers on the X axis represent for calendar days of March 2020

**Table 1:** Demographics, clinical and real time RT-PCR data of the study participants

|  | Patient 1 | Patient 2 | Patient 3 | Patient 4 | Patient 5 | Patient 6 | Patient 7 | Patient 8 |
| --- | --- | --- | --- | --- | --- | --- | --- | --- |
| Age range | 30's | 40's | 20's | 30's | 20's | 20's | 40's | 10's |
| Gender | Male | Male | Male | Male | Female | Female | Female | Male |
| Arriving in Vietnam from abroad (Yes/No) | Yes | Yes | No | No | No | Yes | Yes | Yes |
| Locally acquired infection (Yes/No) | No | No | Yes | Yes | Yes | No | No | No |
| Days from confirmed diagnosis to enrolment | 2 | 1 | 3 | 2 | 3 | 2 | 1 | 0 |
| Days from admission to enrolment | 2 | 1 | 2 | 1 | 0 | 1 | 2 | 0 |
| Duration of stay | 15 | NA | 17 | 18 | 15 | 12 | 13 | 17 |
| Symptomatic (S)/asymptomatic (A) | S | S | S | A | S | A | S | S |
| Laboratory results |  |  |  |  |  |  |  |  |
| White-cell count (×103per µl) | 4.23 | 6.89 | 3.96 | NA | 6.85 | 4.83 | 3.46 | 8.27 |
| Lymphocyte counts (×103per µl) | 0.8 | 0.54 | 1.08 | NA | 2.94 | 2.50 | 0.88 | 2.40 |
| Hemoglobin ((g/dl) | 13.6 | 14.6 | 16.8 | NA | 15.7 | 15.00 | 11.60 | 15.60 |
| Hematocrit (%) | 42.4 | 43.4 | 41.6 | NA | 37.4 | 35.78 | 28.48 | 38.09 |
| Platelet count (per µl) | 140 | 235 | 187 | NA | 414 | 321.00 | 178.00 | 330.00 |
| Glucose (mg/dl) | 125.7 | 112 | 85 | NA | NA | NA | 76.80 | 98.50 |
| Creatinine (mg/dl) | 0.96 | 1.28 | 1.2 | NA | NA | NA | 0.99 | 1.19 |
| Aspartate aminotransferase (U/liter) | 24 | 17 | 22 | NA | NA | NA | 17.40 | 23.30 |
| Alanine aminotransferase (U/liter) | 23 | 16 | 24 | NA | NA | NA | 17.40 | 25.30 |
| Clinical signs/symptoms (Yes/No) |  |  |  |  |  |  |  |  |
| Fever | Y | Y | Y | N | N | N | N | N |
| Cough | N | Y | N | N | N | N | Y | N |
| Rhinorrhea | N | N | Y | N | Y | N | N | Y |
| Fatigue | N | Y | N | N | N | N | N | N |
| Diarrhea | N | N | N | N | N | N | N | Y |
| Sore throat | N | N | Y | N | N | N | Y | N |
| Muscle pain | N | Y | N | N | N | N | Y | N |
| Headache | N | Y | N | N | N | N | N | N |
| Abdominal pain | N | N | N | N | N | N | N | N |
| Lost sense of smell | N | N | N | N | Y | N | N | N |

Information about duration of stay and clinical and laboratory findings are presented in Table 1.

Metagenomics generated a total of 2–4 million reads per sample in 7/8 included NTS. In the remaining sample, ¼ million reads were obtained (Table 2). SARS-CoV-2 were detected in sequence data obtained from all eight RT-PCR positive NTS samples by both IDseq and DISCVR, but not in the NTS sample. One patient presenting with respiratory infection was co-infected with rhinovirus, which was also detected by metagenomics.

**Table 2:**
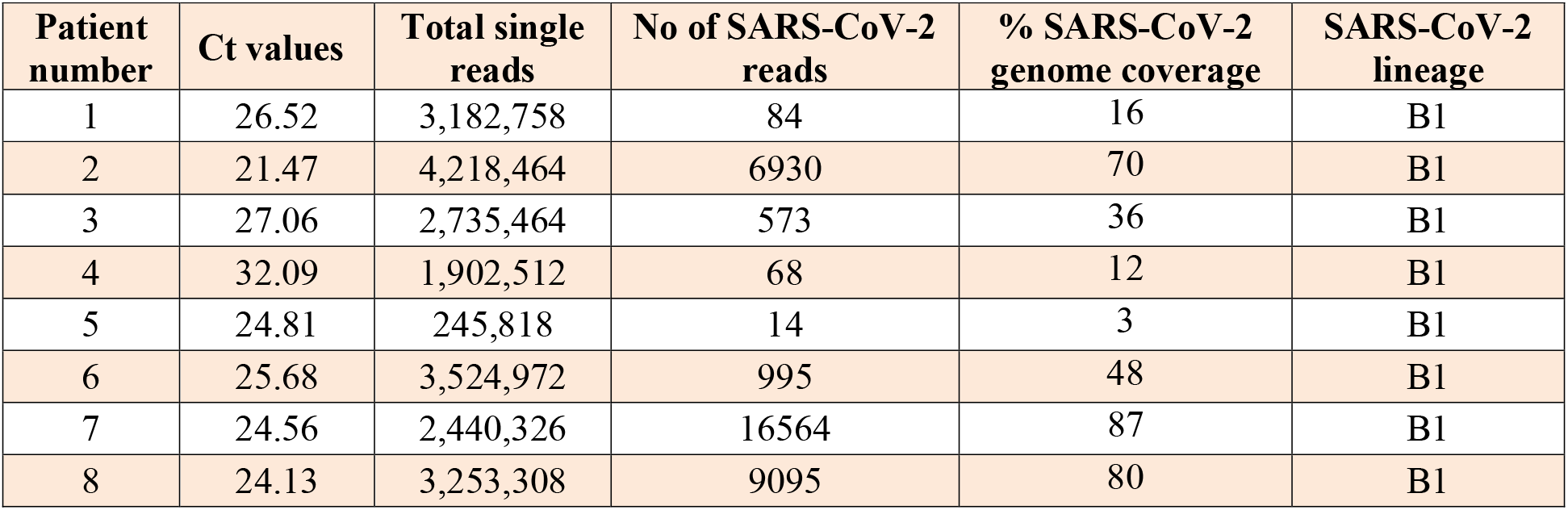
Results of mNGS and lineage assignment SARS-CoV-2 sequences

| Patient number | Ct values | Total single reads | No of SARS-CoV-2 reads | % SARS-CoV-2 genome coverage | SARS-CoV-2 lineage |
| --- | --- | --- | --- | --- | --- |
| 1 | 26.52 | 3,182,758 | 84 | 16 | B1 |
| 2 | 21.47 | 4,218,464 | 6930 | 70 | B1 |
| 3 | 27.06 | 2,735,464 | 573 | 36 | B1 |
| 4 | 32.09 | 1,902,512 | 68 | 12 | B1 |
| 5 | 24.81 | 245,818 | 14 | 3 | B1 |
| 6 | 25.68 | 3,524,972 | 995 | 48 | B1 |
| 7 | 24.56 | 2,440,326 | 16564 | 87 | B1 |
| 8 | 24.13 | 3,253,308 | 9095 | 80 | B1 |

Results of reference-based mapping showed three consensuses had genome coverage of ≥70%, while the remaining five had coverage of <50% (Table 2 and Supplementary Figure 1). Analysis of the obtained consensuses showed all belong to lineage B1. A total of 11 nonsynonymous substitutions were detected in three of the eight obtained consensuses (Supplementary Table 1).

## CONCLUSIONS

The emergence of SARS-CoV-2 emphasizes the continuous unprecedented threat posed by emerging infectious diseases, especially those caused by novel viruses. The diagnosis of respiratory diseases is highly challenging because the responsible pathogens are diverse. In addition, the emergence of novel pathogens further challenges routine diagnosis. Indeed, SARS-CoV-2 initially went undetected by PCR panels targeted at common respiratory viruses [2]. New diagnostic approach is therefore urgently needed to address the ongoing challenge posed by emerging infections.

Here, we demonstrated that when coupled with publically available bioinformatics tools, metagenomics could detect SARS-CoV-2 in RT-PCR positive NTS samples with a wide range of viral loads. The data suggests that metagenomics is a sensitive assay for SARS-CoV-2 diagnosis and detection of co-infection as illustrated by the detection of rhinovirus, in line with a recent report [4], important for clinical management. In addition to providing diagnostic information, the obtained sequences also allows for genetic characterization, and detection of genetic variations in the genomes of the pathogen under investigation. Indeed, using the obtained sequences, we successfully identified that all the Vietnamese viruses included for analysis belonged to lineage B1, which has been found worldwide [11]. In line with a recent report [12], we identify several nonsynonymous substitutions in the obtained genomes SARS-CoV-2. Further research is needed to ascribe the potential consequences that SARS-CoV-2 evolution may have.

Currently, real time RT-PCR is used for screening of suspected cases of SARS-CoV-2 infection [6]. Compared with RT-PCR, metagenomics based on Illumina sequencing technologies remains high cost and low throughput. However, these caveats could be overcome by third generation sequencing technologies such as Oxford Nanopore [13], which warrants further research.

The application of metagenomics for SARS-CoV-2 and respiratory diagnosis would be highly relevant in the near future. This is because SARS-CoV-2 has spread globally, and will likely soon become endemic worldwide. Indeed as of May 21^st^, 2020 nearly 5 million cases have been reported globally. Notably, the vast majority of SARS-CoV-2 infections are asymptomatic or mild, while COVID-19 patients present with signs/symptoms undistinguished with respiratory diseases caused by other viruses [14, 15]. As such rapid identification of the likely cause of hospitalized patients with respiratory infections is essential for clinical management and outbreak response. Under this circumstance, metagenomics is a preferable method because of its ability to detect both known and unknown pathogens presenting in the tested specimens without the need of pathogen specific PCR primers [1, 13].

Our study has some limitations. Only a small number of patients were included for analysis, owing to the nature of a pilot in itself. However during the study period, there were only 14 SARS-CoV-2 confirmed cases reported in our setting, Ho Chi Minh City, Vietnam. As a consequence, we were not able to properly assess the sensitivity and specificity of metagenomics for the diagnosis of COVID-19.

In summary, we show that metagenomics is a sensitive assay for sequence-independent detection of SARS-CoV-2 NTS samples. The ability of metagenomics to detect co-infection and novel pathogens, and generate sequence data for molecular epidemiological investigation makes it an attractive approach for infectious disease diagnosis.

## Data Availability

Provided with in the ms is the essential information. Additional information will be available upon reasonable request.

## ACKNOWLEDGEMENTS

This study was funded by the Wellcome Trust of Great Britain (106680/B/14/Z and 204904/Z/16/Z).

We are indebt to Ms Nguyen Thanh Ngoc, Ms Le Kim Thanh, and the OUCRU IT/CTU/Laboratory Management departments for their support.

We thank the patients for their participations in this study, and the doctors and nurses at HTD Cu Chi Hospital, who cared for the patients and provided the logistic support with the study.

## OUCRU COVID-19 Research Group

**Hospital for Tropical Diseases, Ho Chi Minh City, Vietnam:** Nguyen Van Vinh Chau, Nguyen Thanh Dung, Le Manh Hung, Huynh Thi Loan, Nguyen Thanh Truong, Nguyen Thanh Phong, Dinh Nguyen Huy Man, Nguyen Van Hao, Duong Bich Thuy, Nghiem My Ngoc, Nguyen Phu Huong Lan, Pham Thi Ngoc Thoa, Tran Nguyen Phuong Thao, Tran Thi Lan Phuong, Le Thi Tam Uyen, Tran Thi Thanh Tam, Bui Thi Ton That, Huynh Kim Nhung, Ngo Tan Tai, Tran Nguyen Hoang Tu, Vo Trong Vuong, Dinh Thi Bich Ty, Le Thi Dung, Thai Lam Uyen, Nguyen Thi My Tien, Ho Thi Thu Thao, Nguyen Ngoc Thao, Huynh Ngoc Thien Vuong, Pham Ngoc Phuong Thao, Phan Minh Phuong

**Oxford University Clinical Research Unit, Ho Chi Minh City, Vietnam:** Dong Thi Hoai Tam, Evelyne Kestelyn, Donovan Joseph, Ronald Geskus, Guy Thwaites, H. Rogier van Doorn, Huynh Le Anh Huy, Huynh Ngan Ha, Huynh Xuan Yen, Jennifer Van Nuil, Jeremy Day, Joseph Donovan, Katrina Lawson, Lam Anh Nguyet, Lam Minh Yen, Le Nguyen Truc Nhu, Le Thanh Hoang Nhat, Le Van Tan, Sonia Lewycka Odette, Louise Thwaites, Maia Rabaa, Marc Choisy, Mary Chambers, Motiur Rahman, Ngo Thi Hoa, Nguyen Thanh Thuy Nhien, Nguyen Thi Han Ny, Nguyen Thi Kim Tuyen, Nguyen Thi Phuong Dung, Nguyen Thi Thu Hong, Nguyen Xuan Truong, Phan Nguyen Quoc Khanh, Phung Le Kim Yen, Sophie Yacoub, Thomas Kesteman, Nguyen Thuy Thuong Thuong, Tran Tan Thanh, Tran Tinh Hien, Vu Thi Ty Hang

## ABOUT THE AUTHOR

Dr Le Van Tan is head of Emeging Infections at Oxford Univeristy Clinical Resarh Unit. His research interest includes novel diagnosis and emerging infections.

---

**Supplementary Table 1:** list of non synonymous substitution detected in eight consensuses of the present study

|  | **Nucleotide variation\*** | **Coding Region** | **Amino acid change** | **Detected in GenBank** |
| --- | --- | --- | --- | --- |
| Patient 6 | 17104C>T | nsp13 | H290Y | Yes |
| Patient 7 | 14407C>T | nsp12 | P323L | Yes |
|  | 23402G>A | S | D614G | Yes |
|  | 28881G>A | N | R203K | Yes |
|  | 28882G>A | N | R203K | Yes |
|  | 28883G>C | N | G204R | Yes |
| Patient 8 | 14407C>T | nsp12 | P323L | Yes |
|  | 23402G>A | S | D614G | Yes |
|  | 28881G>A | N | R203K | Yes |
|  | 28882G>A | N | R203K | Yes |
|  | 28883G>C | N | G204R | Yes |

**Note to supplementary Figure 1:** \*compared with reference strain (GenBank accession number NC\_045512.2)

**Supplementary Figure 1:**
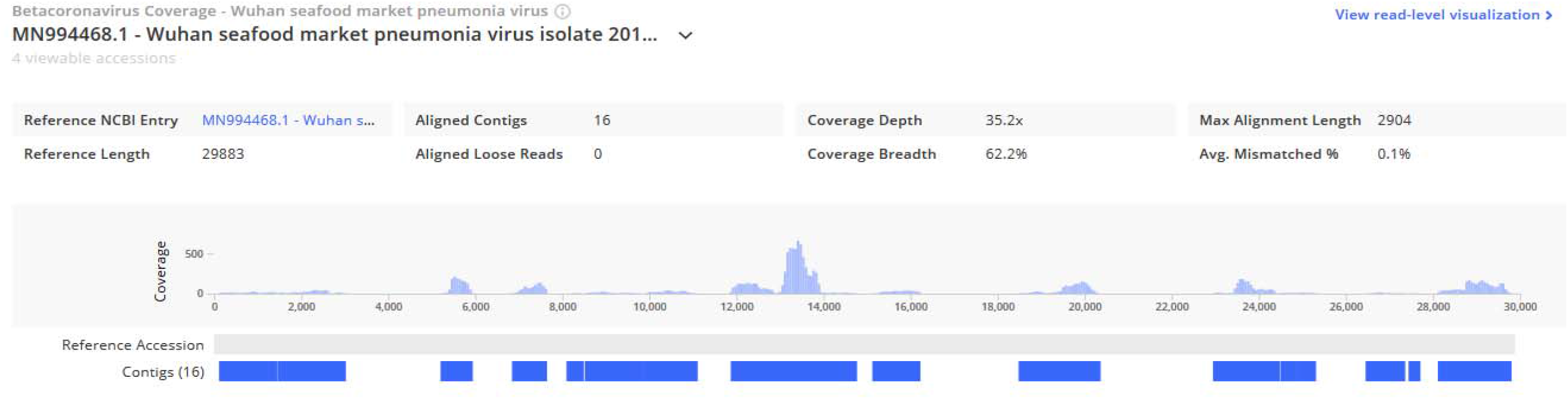
A screen shot showing evidence of SARS-CoV-2 detection in metagenomics data using IDseq pipeline

## Notes

### Competing Interest Statement

The authors have declared no competing interest.

### Author Declarations

Institutional Review Board of the HTD and the Oxford Tropical Research Ethics Committee of the University of Oxford

